# Travel health needs in people visiting friends and relatives: a retrospective analysis of the UK National Travel Health Advice Line, 2019-2025

**DOI:** 10.64898/2026.07.17.26358325

**Authors:** Natalie Elkheir, Sanch Kanagarajah, Dipti Patel

**Affiliations:** Clinical Research Department, Faculty of Infectious and Tropical Diseases, London School of Hygiene & Tropical Medicine; National Travel Health Network & Centre, University College London Hospitals NHS Trust

## Abstract

**Background:** Travellers visiting friends and relatives (VFRs) experience a disproportionate burden of travel-associated infectious diseases, yet little is known about the complexity of pre-travel consultations required to support their care. We compared enquiries relating to VFR travellers and tourists received by the UK National Travel Health Network and Centre (NaTHNaC) specialist Advice Line to identify differences in traveller characteristics, destinations and clinical complexity.

**Methods:** We conducted a retrospective observational study of enquiries to the NaTHNaC Advice Line between 1 January 2019 and 31 December 2025. Enquiries relating to VFR travellers and tourists were compared using descriptive statistics and appropriate statistical tests. Traveller demographics, travel characteristics, destinations and enquiry management were analysed.

**Results:** Of 16,367 enquiries relating to specific travellers, 3,090 (18.9%) concerned VFR travellers and 7,237 (44.2%) concerned tourists. Compared with tourists, VFR travellers were younger (median age 24 vs 52 years, *P*<0.001), more likely to undertake long-stay (8.4% vs 1.8%, *P*<0.001) and last-minute travel (5.0% vs 1.1%, *P*<0.001), and more frequently travelled to the WHO African Region (56.6% vs 29.2%, *P*<0.001) and Eastern Mediterranean Region (12.7% vs 2.8%, *P*<0.001). Pregnancy was substantially more common among VFR travellers (11.6% vs 4.3%, *P*<0.001). Enquiries concerning VFR travellers were more likely to require a call-back (16.1% vs 13.8%, *P*=0.012) and escalation to a specialist doctor (13.1% vs 10.5%, *P*<0.001), indicating greater consultation complexity. General practice generated a higher proportion of VFR-related enquiries than tourist enquiries (69.5% vs 63.9%, *P*<0.001).

**Conclusions:** VFR travellers generate disproportionately complex pre-travel consultations characterised by higher rates of specialist escalation, distinct travel patterns and travel to destinations associated with the greatest burden of imported infectious diseases. These findings highlight the importance of specialist travel medicine support for healthcare professionals managing VFR travellers and reinforce the need for equitable access to timely, high-quality pre-travel healthcare for this high-risk population.

## Background

International travel is associated with the importation of a wide range of infectious diseases, many of which are preventable through appropriate pre-travel risk assessment, vaccination, malaria chemoprophylaxis and health advice.^1-3^ Despite these preventive measures, imported infections such as malaria, enteric fever and hepatitis A continue to cause substantial morbidity among UK travellers. Travellers visiting friends and relatives (VFRs), typically first- and second-generation migrants returning to countries of origin or heritage, experience a disproportionate burden of these infections.^2,4^ In the UK, VFR travellers account for the majority of imported malaria cases and most imported enteric fever and hepatitis A cases, with many infections occurring in individuals who did not receive recommended pre-travel interventions such as malaria chemoprophylaxis or vaccinations.^5-7^

Compared with tourists, VFR travellers are more likely to undertake longer journeys, stay within local communities, travel at short notice, and have lower uptake of pre-travel health advice, vaccination and malaria chemoprophylaxis.^8-10^ Consequently, VFR travellers may present to travel health providers with complex needs that require specialist advice beyond routine travel clinic consultations.

The National Travel Health Network and Centre (NaTHNaC) provides the UK’s specialist travel health advice service, supporting healthcare professionals managing complex pre-travel enquiries through an expert telephone advice line.^11^ Previous analyses of advice line activity have described the characteristics of enquiries received and demonstrated the value of the service as a national source of specialist travel medicine expertise.^12-15^ However, differences in the nature of enquiries relating to VFR travellers and leisure travellers (tourists) have not been explored.

The aim of this study was to compare the characteristics of NaTHNaC advice line enquiries relating to VFR travellers and tourists received between 2019 and 2025, including traveller demographics, destinations, and reasons for consultation. By identifying differences in the complexity and nature of enquiries between these groups, we sought to inform travel health practice, education, and future service provision.

## Methods

### Study design and setting

We conducted a retrospective observational study of enquiries received by the NaTHNaC Advice Line between 1 January 2019 and 31 December 2025. The NaTHNaC Advice Line is a national specialist service providing evidence-based travel health advice to healthcare professionals managing complex pre-travel consultations in the United Kingdom. Travel health providers (clinicians, travel health nurses, pharmacists and doctors) phone the enquiry line during designated time slots throughout the week, to speak with a highly experienced NaTHNaC specialist nurse. Enquiries relate to a broad range of travel health issues, including vaccination, malaria prevention, complex medical conditions, pregnancy, immunocompromise, and itinerary-specific risk assessment. NaTHNaC is the UK’s national clinical travel health service, providing specialist evidence-based advice to healthcare professionals managing complex travel health enquiries. During each consultation, responding clinicians record enquiry details using a standardised electronic proforma completed as part of routine clinical practice.

### Data collection

Advice line records were extracted from the NaTHNaC enquiry database. Each record represents a single enquiry relating to one or more travellers. Information routinely collected includes caller characteristics, traveller demographics, travel destination, purpose of travel, relevant medical history, immunocompromise, medications, enquiry topic and advice provided.

### Study population

For this study, we included enquiries relating to travellers whose purpose of travel was recorded as either visiting friends and relatives (VFR) or tourism. Enquiries relating to other travel purposes (including business, expatriate assignments, pilgrimage, aid work and travel for healthcare) and those with missing or unknown travel purpose were excluded from the comparative analyses. The purpose of travel is recorded as a predefined variable within the advice line proforma.

### Variables

Variables extracted included: date of enquiry; traveller age and sex; destination country or region; long-stay or last-minute travel; presence of special health needs; immunocompromised status; relevant medical history and current medications; and the principal reason for the enquiry, including vaccine-related, malaria-related and other general travel health advice.

Where enquiries related to multiple travellers, data were extracted for the traveller(s) included in the enquiry record.

### Statistical analysis

Characteristics of enquiries relating to VFR travellers and tourists were summarised using descriptive statistics. Categorical variables were compared using a χ^2^ test, while continuous variables were compared using Student’s *t*-test if normally distributed or the Mann–Whitney *U* test if not. All tests were two-sided, with *P*<0.05 considered statistically significant. Statistical analyses were performed using STATA version 18.0.

### Ethics

This study analysed routinely collected service data that did not include personal identifiable information. The project was undertaken as a retrospective evaluation of a national specialist travel health service and so formal ethics committee review was not required.

## Results

### Study population

Between 1 January 2019 and 31 December 2025, the NaTHNaC Advice Line received 18,663 enquiries. After excluding 2,296 enquiries that did not relate to a specific traveller or group, the purpose of travel for the remaining 16,367 enquiries is depicted in **table 1**.

**Table 1.** Purpose of travel for 16,367 telephone enquiries to the NaTHNaC advice line, 2019-2025.

| Purpose of travel | Number of travellers (%) |
| --- | --- |
| Tourism | 7,237 (44.2%) |
| Visiting friends and relatives | 3,090 (18.9%) |
| Not known | 1,421 (8.7%) |
| Multiple | 877 (5.4%) |
| Cruise | 838 (5.1%) |
| Other | 706 (4.3%) |
| Business | 637 (4.1%) |
| Backpacker | 457 (2.8%) |
| School | 370 (2.3%) |
| Aid worker | 248 (1.5%) |
| Pilgrim | 189 (1.2%) |
| Expat | 162 (1%) |
| Healthcare worker | 46 (0.3%) |
| Travel to altitude | 30 (0.2%) |
| Gap year | 19 (0.1%) |
| Medical tourist | 4 (<0.1%) |

Of all 16,367 enquiries related to specific travellers, 3,090 (18.9%) concerned VFR travellers and 7,237 (44.2%) concerned tourists, who were selected as the comparator group for the rest of the analysis.

### Characteristics of calls

Primary care clinicians were the most frequent callers to the advice line for both traveller groups but accounted for a greater proportion of enquiries relating to VFR travellers than tourists (70% vs 64%, p<0.001). Conversely, pharmacies generated a lower proportion of VFR enquiries than tourist enquiries (20% vs 26%, p<0.001). Approximately two-thirds of enquiries originated from registered yellow fever vaccination centres, with no significant difference between traveller groups. Most enquiries related to a single traveller in both groups. Caller characteristics are summarised in **table 2**. Advice line management differed between traveller groups. Calls about VFR travellers were more likely to require a call back (16% versus 14%, p=0.012) and/or escalation to a doctor (13% vs 11% p<0.001), suggesting more complex enquiries. Approximately half of enquiries in both groups were managed using guidance from the NaTHNaC website, while use of the UK ‘Green Book’ Immunisation against infectious disease and NaTHNaC protocols was similar between groups.

**Table 2.** NaTHNaC advice line call characteristics relating to tourists and VFR travellers, 2019-2025.

|  | **Tourists**<br>(n calls = 7,237) | **VFR travellers**<br>(n calls = 3,090) | P-value* |
| --- | --- | --- | --- |
| **Workplace of caller** |  |  |  |
| GP | 4,626 (63.9%) | 2,149 (69.5%) | <0.001 |
| Pharmacy | 1,891 (26.1%) | 622 (20.1%) |  |
| Private travel clinic | 547 (7.6%) | 257 (8.3%) |  |
| Other | 133 (1.8%) | 40 (1.3%) |  |
| Occupational health | 18 (0.2%) | 4 (0.1%) |  |
| No data | 15 (0.2%) | 10 (0.3%) |  |
| Military | 7 (0.1%) | 8 (0.3%) |  |
| **Yellow fever vaccination centre** | 4,619 (63.8%) | 1,908 (61.7%) | 0.133 |
| **Number of travellers the call is about** |  |  |  |
| 1 | 6,408 (88.5%) | 2,640 (85.4%) | <0.001 |
| 2 | 743 (10.3%) | 338 (10.9%) |  |
| 3 | 52 (0.7%) | 85 (2.8%) |  |
| **Advice†** |  |  |  |
| Green book | 586 (8.1%) | 197 (6.4%) | <0.001 |
| NaTHNaC protocols | 36 (0.5%) | 9 (0.3%) |  |
| NaTHNaC website | 3,769 (52.1%) | 1,505 (48.7%) |  |
| Multiple | 2,486 (34.4%) | 1,181 (38.2%) |  |
| Other guidelines | 169 (2.3%) | 82 (2.7%) |  |
| Missing/no data | 47 (0.6%) | 35 (1.1%) |  |
| Call back required | 1,002 (13.8%) | 497 (16.1%) | 0.012 |
| Escalation to doctor required | 763 (10.5%) | 405 (13.1%) | <0.001 |

\* P value refers to statistical tests assessing differences between calls about tourists and VFR travellers ( $\chi^2$ tests for categorical variables).
† Multiple types of advice could be provided to enquirers and so totals can sum to >100%.

### Travel destinations

Marked differences were observed in travel destination (**table 3**). Over half of VFR enquiries related to travel to the WHO African Region (56.6%), compared with fewer than one-third of tourist enquiries (29.2%) (*p*<0.001). VFR travellers were also more likely to travel to the Eastern Mediterranean Region (12.7% vs 2.8%) and the Western Pacific Region (11.7% vs 8.9%). In contrast, tourist enquiries more frequently involved travel to the Americas (17.7% vs 6.6%) and South-East Asia (9.3% vs 4.6%), while itineraries involving multiple WHO regions were considerably more common among tourists (27.5% vs 4.7%). Kenya, Tanzania and Brazil were the three most visited countries generating enquiries for tourists. Whereas, Ghana, Nigeria and Pakistan were the three most visited countries generating enquiries for VFR travellers.

**Table 3.** Traveller destinations by tourism and VFR travel reported to NaTHNaC advice line, 2019 -2025.

|  | Tourists | VFR travellers | P-value* |
| --- | --- | --- | --- |
|  | (n calls = 7,237) | (n calls = 3,090) |  |
| Traveller destination<br>(WHO region) |  |  |  |
| Africa | 2,113 (29.2%) | 1,750 (56.6%) | <0.001 |
| Multiple | 1,988 (27.5%) | 146 (4.7%) |  |
| Americas | 1,281 (17.7%) | 205 (6.6%) |  |
| South East Asia | 670 (9.3%) | 142 (4.6%) |  |
| Western Pacific | 645 (8.9%) | 360 (11.7%) |  |
| Eastern Mediterranean | 204 (2.8%) | 392 (12.7%) |  |
| Europe | 143 (2%) | 49 (1.6%) |  |
| Missing | 112 (1.5%) | 45 (1.5%) |  |
| Worldwide | 81 (1.1%) | 1 (<0.1%) |  |
| Top 10 countries |  |  |  |
|  | Kenya, n = 1,284 | Ghana, n = 553 | <0.0001 |
|  | Tanzania, n =702 | Nigeria, n = 352 |  |
|  | Brazil, n = 619 | Pakistan, n = 294 |  |
|  | Peru, n = 526 | Uganda, n = 233 |  |
|  | Thailand, n = 486 | Indonesia, n = 221 |  |
|  | South Africa, n = 461 | Kenya, n = 148 |  |
|  | Indonesia, n = 441 | Brazil, n = 106 |  |
|  | Argentina, n = 341 | Sierra Leone, n = 63 |  |
|  | Vietnam, n = 272 | Bangladesh, n = 58 |  |
|  | Gambia, n = 246 | Philippines, n = 56 |  |
| *P value refers to statistical tests assessing differences between calls about tourists and VFR travellers (χ² tests for categorical variables). |  |  |  |

**Table 4.**
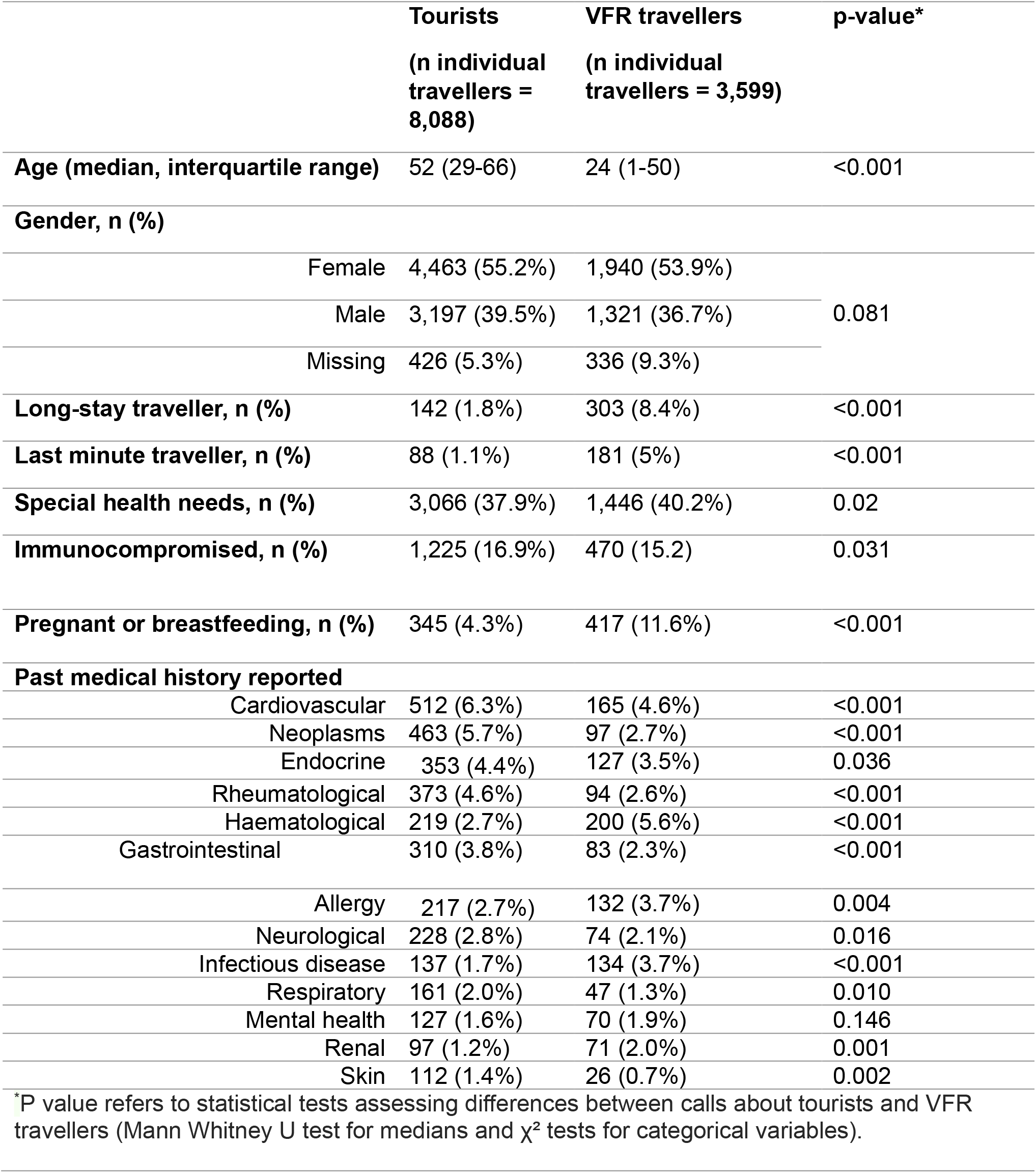
Traveller characteristics by tourism and VFR travel of enquiries reported to NaTHNaC advice line, 2019 -2025.

### Traveller characteristics

VFR travellers were substantially younger than tourists (median age 24 years [IQR 1–50] versus 52 years [IQR 29–66], *P*<0.001) (table 3). There was no significant difference in traveller gender between groups. However, VFR travellers were more likely to be undertaking long-stay travel (8.4% vs 1.8%, *P*<0.001) and last-minute travel (5% vs 1.1%, *P*<0.001). Special health needs were also slightly more common among VFR travellers than tourists (40.2% vs 37.9%, *P*=0.02). A much higher proportion of VFR travellers were pregnant compared to tourists (11.6% versus 4.3%, p<0.001). There were significant differences in past medical history reported between groups. Tourists had higher proportions of cardiovascular disease, neoplasms, endocrine disease, rheumatological disease, gastrointestinal disease, neurological disease, respiratory disease and skin conditions. Whereas VFR travellers had higher proportions of haematological disease, allergy, infectious disease, mental health conditions and renal disease.

## Discussion

This study compared specialist travel health enquiries relating to VFR travellers to those related to tourists, managed through a national travel medicine advice service. We found that almost one in five enquiries concerned VFR travellers and that these enquiries differed substantially from those relating to tourists in terms of traveller demographics, destinations, travel characteristics and clinical complexity. Importantly, enquiries concerning VFR travellers were significantly more likely to require a call-back or escalation to a specialist doctor, suggesting that these consultations pose greater challenges for travel health providers than those involving tourists.

Our findings support the large body of literature demonstrating that VFR travellers experience a disproportionate burden of travel-associated infectious diseases. GeoSentinel surveillance has consistently shown that VFR travellers are overrepresented among cases of imported malaria, enteric fever and viral hepatitis, and are more likely to require hospitalisation than tourists travelling to the same destinations.^2,3^ These observations are mirrored in UK surveillance, where VFR travellers account for the majority of imported malaria cases and a substantial proportion of imported enteric fever and hepatitis A despite representing a minority of international travellers.^5,7^ The predominance of VFR enquiries in our study relating to travel to sub-Saharan Africa and South Asia, particularly Ghana, Nigeria and Pakistan, closely reflects the epidemiology of imported infections reported through UK surveillance systems and supports the representativeness of the Advice Line population.^5,7^

Consistent with previous studies, VFR travellers were substantially younger than tourists and were significantly more likely to undertake long-stay and last-minute travel.^8,9^ Both characteristics reduce opportunities to complete recommended vaccination schedules and optimise malaria chemoprophylaxis while increasing cumulative exposure to infectious diseases. Longer stays within local communities, rather than tourist accommodation, also increase opportunities for mosquito exposure, food- and water-borne infections and close contact with local populations, factors that have been proposed as key contributors to the higher burden of imported disease observed among VFR travellers.^4,8^

We also observed important differences in clinical characteristics between traveller groups. Pregnancy was almost three times more common among VFR travellers, while special health needs were also slightly more frequent. These factors substantially complicate travel risk assessment, particularly for travel to malaria-endemic regions where clinicians must balance the maternal risks of malaria against the safety, efficacy and suitability of available chemoprophylaxis. Although tourists more frequently reported chronic conditions such as cardiovascular disease, malignancy and rheumatological disorders, these largely reflected their older age profile and are generally supported by well-established travel medicine guidance. Leisure travellers can chose where they travel and so those with more complex vulnerability (including comorbidities and pregnancy) have the option of choosing their destinations based on their individual health needs. By comparison, consultations involving VFR travellers often required consideration of multiple interacting factors—including destination-specific disease epidemiology, pregnancy, travel timing and prolonged exposure—which may explain the greater requirement for specialist medical input observed in our study.

Interestingly, a greater proportion of VFR-related enquiries originated from general practice, whereas pharmacies generated proportionally fewer enquiries than for tourists. This may reflect differing healthcare utilisation or the greater complexity of VFR consultations and the broader medical history often available within primary care.

Lower uptake of pre-travel advice among VFR travellers has been consistently reported internationally and is widely recognised as a major contributor to preventable travel-associated illness.^4,8,9^ Previous studies have demonstrated that VFR travellers are less likely to seek pre-travel healthcare or receive recommended malaria chemoprophylaxis and vaccinations than tourists. These disparities should not be viewed solely as individual behavioural choices but rather as reflecting multiple interacting barriers, including financial costs, limited access to specialist travel services, time pressures associated with urgent family travel, language and cultural factors, competing priorities and lower perceived susceptibility when returning to familiar environments.^4,8^ Addressing these barriers will require culturally appropriate approaches to travel health promotion alongside improved access to affordable pre-travel services within primary care and community settings.

An important implication of our findings is the continued value of national specialist travel medicine services. Approximately half of all enquiries utilised NaTHNaC’s online guidance, yet many consultations required reference to multiple guidance sources, call-backs or escalation to a specialist clinician. This suggests that written guidance alone cannot address every complex travel scenario and that expert consultation remains an important component of high-quality travel medicine practice. Rather than duplicating online resources, specialist advice services appear to provide complementary support where evidence is limited, risks are finely balanced or individual patient circumstances require nuanced interpretation of existing recommendations.

The principal strengths of this study include its large national dataset collected over seven years and the use of routinely recorded clinical information from a specialist service accessed by healthcare professionals across primary care, pharmacy and specialist travel clinics. Unlike surveillance studies describing imported disease after travel, this study provides a unique perspective on the complexity of pre-travel risk assessment and the circumstances in which clinicians seek specialist support.

Several limitations should be acknowledged. First, the study represents enquiries made to a specialist advice service and therefore reflects a selected population of more complex consultations rather than all UK travellers. Secondly, the analysis relied on routinely collected service data and some variables were incompletely recorded. Thirdly, travel purpose was based on clinician-recorded information and some travellers with multiple reasons for travel may have been misclassified. Finally, because this was an observational study, we cannot determine whether specialist advice altered subsequent travel health outcomes or reduced travel-associated disease.

In conclusion, VFR travellers generate disproportionately complex pre-travel consultations characterised by higher rates of specialist escalation, distinct travel patterns and destinations associated with the greatest burden of imported infectious diseases. While previous studies have highlighted the increased morbidity experienced by VFR travellers after travel, our findings demonstrate that this complexity is already evident during pre-travel assessment. Strengthening access to specialist travel medicine advice, alongside interventions that improve equitable access to pre-travel healthcare for migrant communities, may help reduce preventable travel-associated infections and support clinicians managing this high-risk group.

## Data Availability

All data produced in the present study are available upon reasonable request to the authors.

